# A Syndromic Surveillance Tool to Detect Anomalous Clusters of COVID-19 Symptoms in the United States

**DOI:** 10.1101/2020.08.18.20177295

**Authors:** Amparo Güemes, Soumyajit Ray, Khaled Aboumerhi, Michael R. Desjardins, Anton Kvit, Anne E Corrigan, Brendan Fries, Timothy Shields, Robert D Stevens, Frank C Curriero, Ralph Etienne-Cummings

## Abstract

Coronavirus SARS-COV-2 infections continue to spread across the world, yet effective large-scale disease detection and prediction remain limited. COVID Control: A Johns Hopkins University Study, is a novel syndromic surveillance approach, which collects body temperature and COVID-like illness (CLI) symptoms across the US using a smartphone app and applies spatio-temporal clustering techniques and cross-correlation analysis to create maps of abnormal symptomatology incidence that are made publicly available. The results of the cross-correlation analysis identify optimal temporal lags between symptoms and a range of COVID-19 outcomes, with new taste/smell loss showing the highest correlations. We also identified temporal clusters of change in taste/smell entries and confirmed COVID-19 incidence in Baltimore City and County. Further, we utilized an extended simulated dataset to showcase our analytics in Maryland. The resulting clusters can serve as indicators of emerging COVID-19 outbreaks, and support syndromic surveillance as an early warning system for disease prevention and control.

## Introduction

Despite progress in the fight against COVID-19, the pandemic remains the most immediate threat to human health and society. Substantial efforts have been deployed to investigate this disease, yet our knowledge of its origin, progress, and biological mechanisms remains limited. An effective vaccine and treatment are imperative to reducing disease burden, but until either is achieved, efforts must be focused on prevention of disease transmission, which necessitates robust disease surveillance and response. Current approaches implemented to manage the spread of the infection could be significantly improved by clearly discriminating between high and low risk population areas and time periods.

Metrics currently used to describe the progress of COVID-19 are based primarily on cases confirmed by laboratory tests, numbers of patients hospitalized and in intensive care, and counts of the number of deaths^1,2^. The data for each of these metrics, however, is temporally inconsistent and often delayed due to incubation time of the virus (2-14 days), the time from the onset of symptoms until clinical care is obtained, the time from test to confirmed test result, and, in the more severe cases, the time from hospital admission to death.^2,3^ It is estimated existing epidemiological accounts of COVID-19, when measured in counts of confirmed cases or deaths, provides a snapshot of infections acquired 2-4 weeks previously.^4^ To further complicate matters, the information around each of these sources of error has varied dramatically since the start of the pandemic in March 2020. These metrics might be nevertheless highly informative if it is assumed that all infected individuals seek medical attention and are thus incorporated into the case statistics. These metrics, however, do not consider the population that is infected but is completely asymptomatic (40%-50% in many studies),^5^ or those who prefer not to seek medical care despite having symptoms. Furthermore, when hospitalizations do occur, they often happen days after contracting the disease, when symptoms begin to manifest or worsen. Consequently, it is essential to complement these disease monitoring systems with syndromic surveillance systems that allow a more extensive and timely evaluation of the population.

Strategies for monitoring and predicting influenza-like illnesses (ILI) are essential when implementing a surveillance system for COVID-19^6^, because many signs and symptoms of COVID-19 are nonspecific and can be indicative of other illnesses such as influenza, Lyme Disease, and the common cold. However, there are certain symptoms that are more specific to COVID-19 and may be indicative of the prognosis of the disease. Anosmia (loss of smell) is considered a common early symptom of COVID-19, and based on new findings it may be a predictor of a less severe infection being less likely to require hospitalization.^7^ Other signs such as skin rashes or loss of color in the fingers or toes may also be prognostic indicators, as confirmed by the results of a recent study that classifies skin manifestations into five patterns associated with a specific prognosis of COVID-19 infection.^8^ The CDC has one of the most comprehensive monitoring systems for ILI. One of the sources for data collection consists of statistics on patients presenting flu-like symptoms to healthcare providers through a national network called ILInet. In more detail, 2,600 outpatient healthcare providers nationwide report to the CDC weekly on the percentage of patients diagnosed with influenza out of the total number of patients seen. Presentation of fever (temperature 100° F [37.8° C] or more) and cough and / or sore throat without a known cause other than the flu are the requirements to be diagnosed as ILI. With the data collected in ILINet, the CDC generates a measure of ILI activity at the national, regional and state level.

The epidemiology centers that contribute every year with real-time probabilistic forecasts for pandemic and seasonal influenza activity make up the so-called “network” of Centers of Excellence for Influenza Forecasting of the CDC. One of these institutions is the Delphi Research Group from Carnegie Mellon University. This research group has recently focused their efforts on monitoring and predicting COVID-19, and has developed the COVIDcast system that gathers aggregated data from different sources by collaborating with multiple partners to visually display predictions of COVID-19 activity levels and prevalence in the United States.^9^ The data includes, among others, Google search statistics for COVID-related topics, and short surveys where Facebook and Google users anonymously report whether they know someone or if they themselves present few CLI, including fever, cough, shortness of breath, or difficulty breathing.

Another syndromic study that has emerged in recent months in the United Kingdom and now in the U.S. is the COVID Symptom Study^10^. This is a mobile application in which registered users fill in daily questions about their medical history and the presentation of numerous symptoms of COVID-19. This research aims to help scientists better understand the symptoms of COVID-19,^11^ and in the future, could be used to track the spread of this virus, and identify high-risk areas in the country. Finally, Kinsa, a company developing and distributing smart thermometers, is now collecting the body temperature of its customers to create maps showing the areas where fever levels are abnormally high, compared to the levels expected for the time of year, which may be early indicators of the spread of COVID-19.^12^ Additionally, they create estimates of the activity trend using methods previously described by Dalziel et al.38 that would be expected under normal influenza conditions, which allows them to identify activity levels that are higher than this ‘normality’.^13^

While these are examples of systems that have been implemented to track or map COVID-19 cases or symptoms, there is still an unmet need for a tool for the detection of anomalous outbreaks of CLI symptoms that is spatially accurate and continually updated. In an effort to overcome these challenges and support more selective mitigation strategies, we developed a syndromic surveillance system that is comprised of an app to gather CLI data, coupled with space-time analytics to identify hotspots of anomalous CLI symptoms and, in turn, identify potential clusters of COVID-19. Our tool meets the following characteristics: 1) allows accurate mapping of CLI in space and time; 2) reflects in quasi-real time the health status of the sampled population; 3) is broadly accessible and easy to use, and 4) collected data is intuitively visualizable and widely disseminated (e.g. via a dashboard). Detecting disease clusters in space and time is an exploratory approach in infectious disease surveillance^14–16^ which identifies geographical locations or regions where the observed number of cases or symptoms exceeds the expected number of cases given baseline conditions.^17^ Here, we report syndromic surveillance results obtained using purely temporal and space-time cluster detection approaches. We also detected space-time clusters derived from a simulated dataset of over 800,000 entries in Maryland to showcase our analytics when our app has an increased userbase.

## Results

The app was launched on 25 April 2020, and since that date has been installed by over 11,000 unique users with more than 72,000 data entries in 1,019 counties across all states in the US (data on July 25, 2020). Age distribution of users was bimodal with a peak in the 18-25 age bin and a second peak at 51-55 years (Figure 1). Sex distribution is currently not relevant for the study. The majority of users (84.7%) entered data for a maximum of 2 weeks (Figure 2).

**Figure 1.**
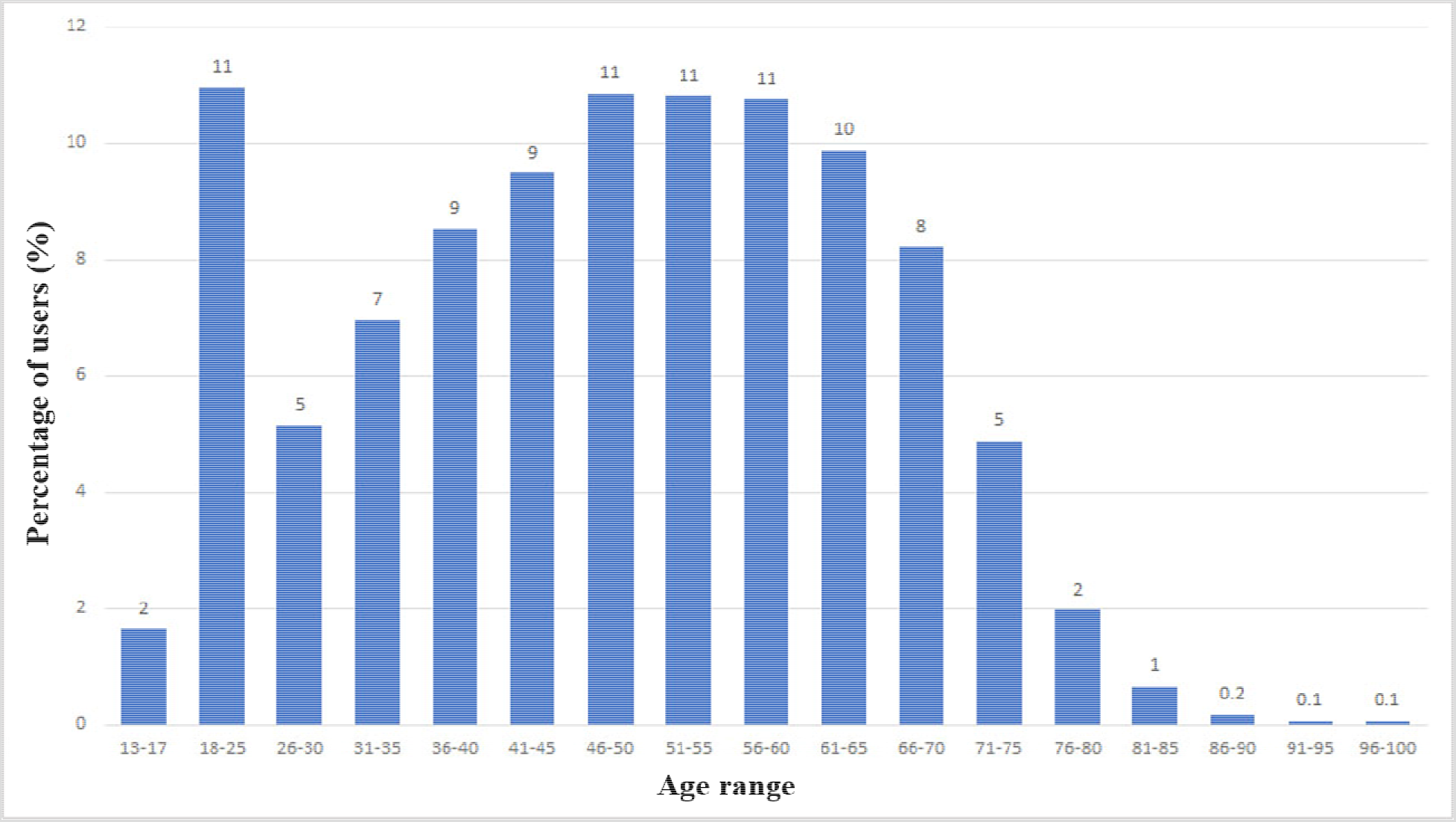
Demographics of app users up to July 25, 2020 deri

**Figure 2.**
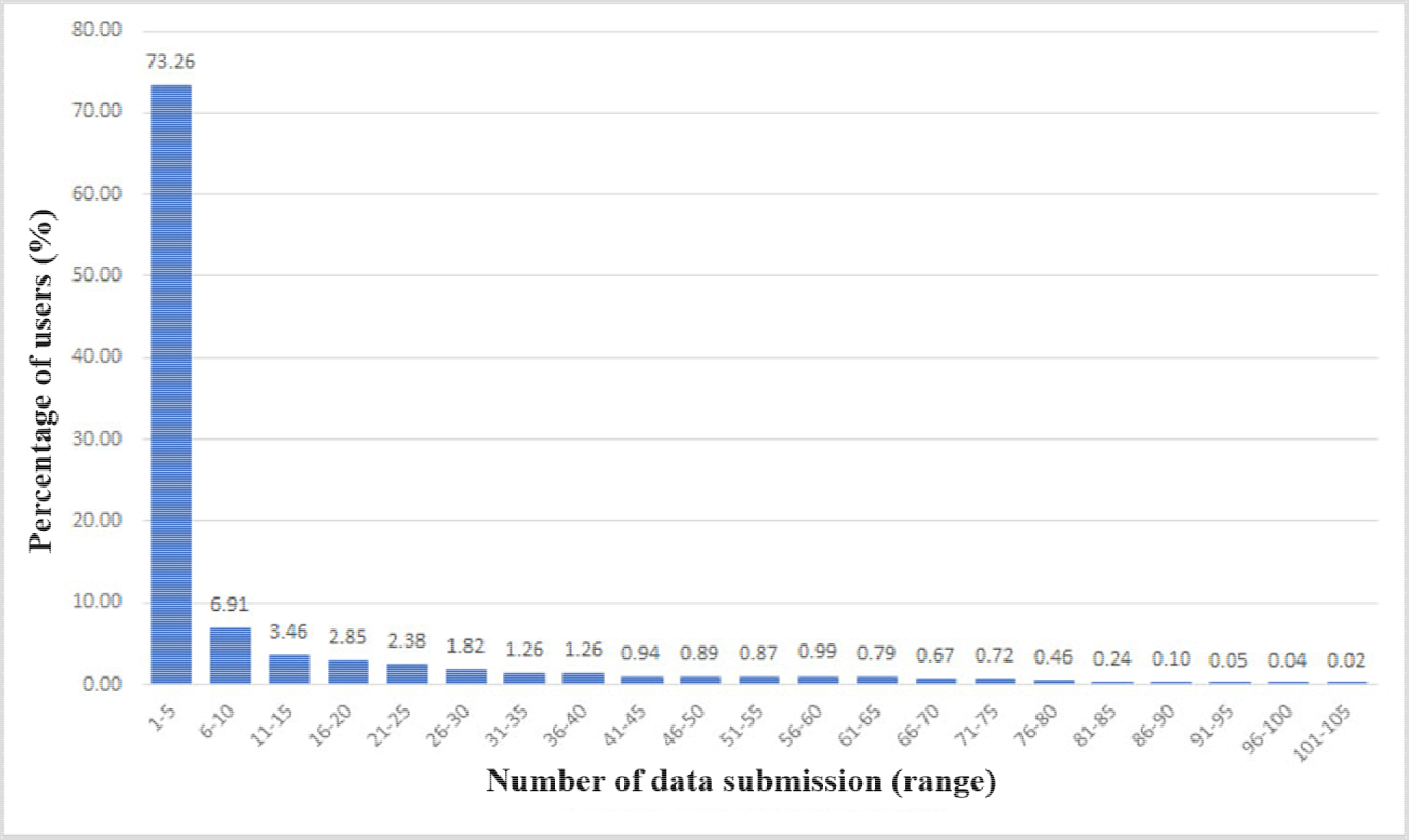
Distribution of number of submissions per user up July 25, 2020

The time series for the number of daily data entries of each symptom from app launch to July 25, 2020 is shown in Figure 3.

**Figure 3.**
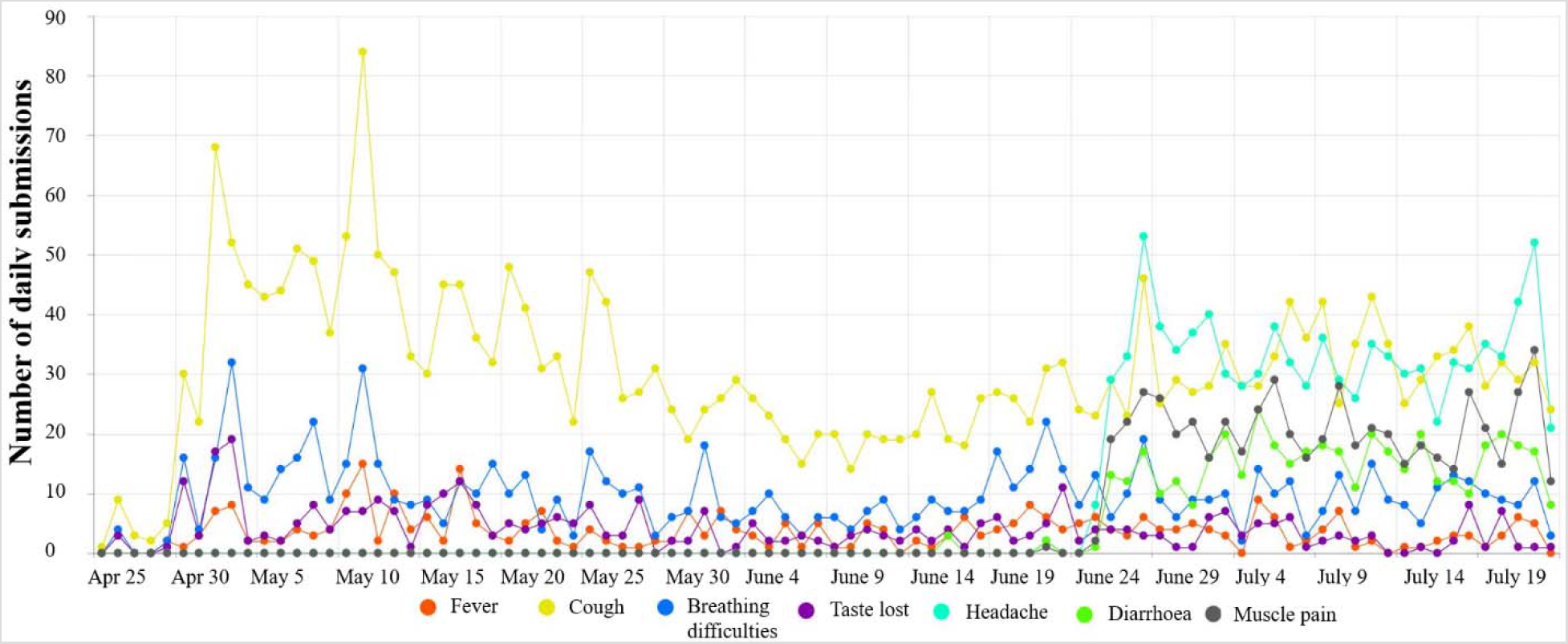
Prevalence of self-reported symptoms up to July 25, 2020

Figure 4 provides the results of the retrospective cluster detection analysis in Baltimore County and City from between April 26^th^, 2020 and June 10^th^, 2020. As usership increased, the number of reported CLI symptoms also increased. A statistically significant cluster of symptoms was first detected in the northern region of Baltimore City (Cluster 1), with a duration of 5 days (Apr. 27^th^-May 1^st^). Cluster 1 includes 20 observed CLI symptom cases, with an expected count of 1.8. Cluster 2 was detected on May 18^th^ southeastern Baltimore County with 6 observed and 0.1 expected symptoms. Cluster 3 was detected in the southwestern region of Baltimore County on May 3^rd^, with a duration of 6 days. Cluster 3 includes 24 observed and 8.3 expected symptom cases. A fourth cluster was detected (Cluster 4) on May 9^th^, with 5 observed and 0.2 expected CLI symptom cases in western Baltimore County and portions of western Baltimore City. When evaluating the relationship between app-recorded symptoms and confirmed COVID-19 cases in Maryland, the strongest predictor was new loss of taste/smell (correlation coefficient of 0.65; p < 0.01) at a 5-day lag.

**Figure 4.**
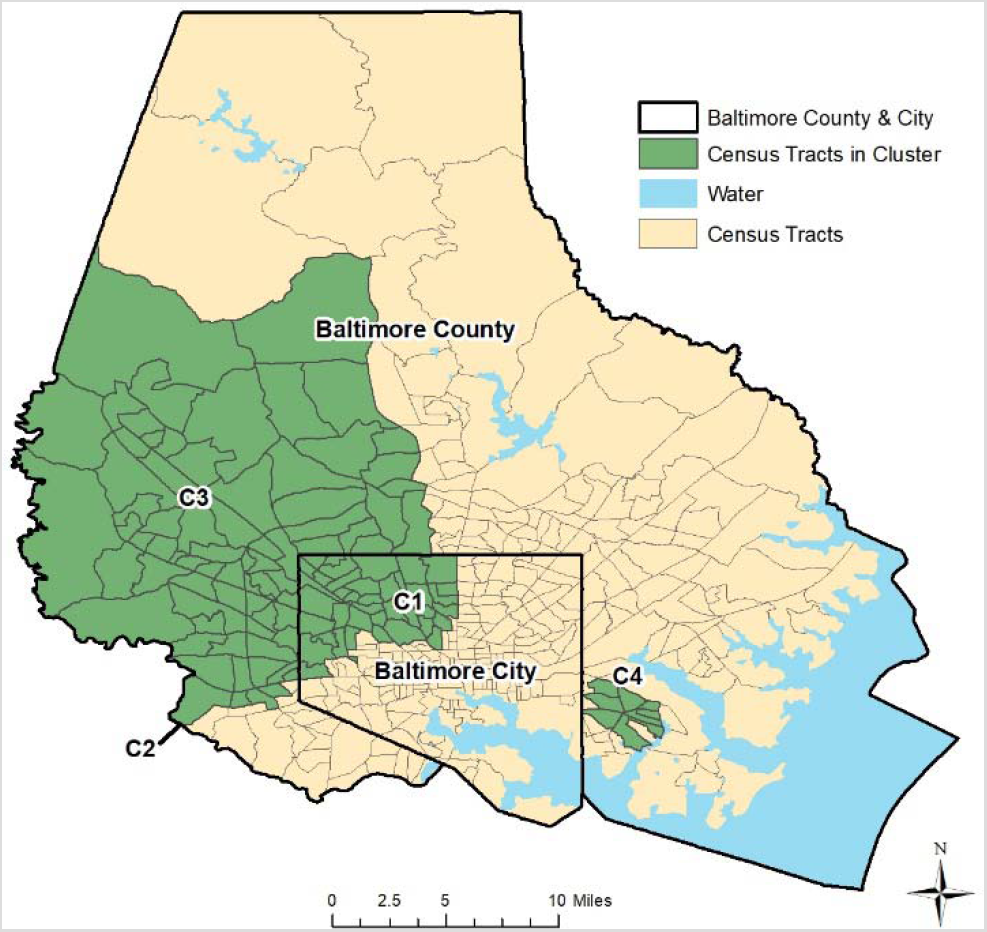
Space-time clusters of COVID Control Symptoms in Baltimore County & City, Maryland

Simulated cases were generated for Maryland state from a bivariate Gaussian distribution in space and a lognormal distribution in time. Two clusters, varying both in the spatial and temporal dynamics were randomly chosen. Based on the combined spatiotemporal case distribution, a genetic algorithm was used to fit generated user profiles to the cases counts. These user profiles were then used to generate symptom information using the actually reported prevalence of each symptom, along with random noise to simulate self-reported symptom information. Using our simulated dataset of 836,721 entries (91,674 symptomatic entries – cases; and 745,047 asymptomatic entries – controls), we detected space-time clusters in 16 of 24 Maryland counties between May 24^th^ and June 23^rd^, 2020. Figures 5 and 6 provide the results of the daily space-time cluster detection analysis on June 6^th^ and June 23^rd^, respectively (i.e. first day and last day of analysis), On June 6^th^, 15 of Maryland’s 24 counties contained at least one space-time cluster; two in the western-most counties (Garrett and Allegany) and 13 in the central and north-eastern regions of the state. One June 23^rd^, 14 counties contained at least one significant space-time cluster; while there were no longer clusters in the western counties and Talbot County contained its first since the start of the analysis on June 6^th^. The clusters on June 23^rd^ also covered less area in Maryland, suggesting an overall decrease in CLI-symptom activity/entries. Table 1 summarizes the daily results by presenting the number of times a Maryland county contained a cluster; and the dates a cluster was first and last identified in each respective county. Twelve of the counties contained at least one cluster of CLI-like symptoms between June 6^th^ and June 23^rd^ (i.e. 22 days); and 8 counties never contained a cluster during the 18-day period of analysis. The analytical results present an example of how our tool can be used to detect anomalous clusters of CLI symptoms, which will likely grow as userbase increases.

**Figure 5.**
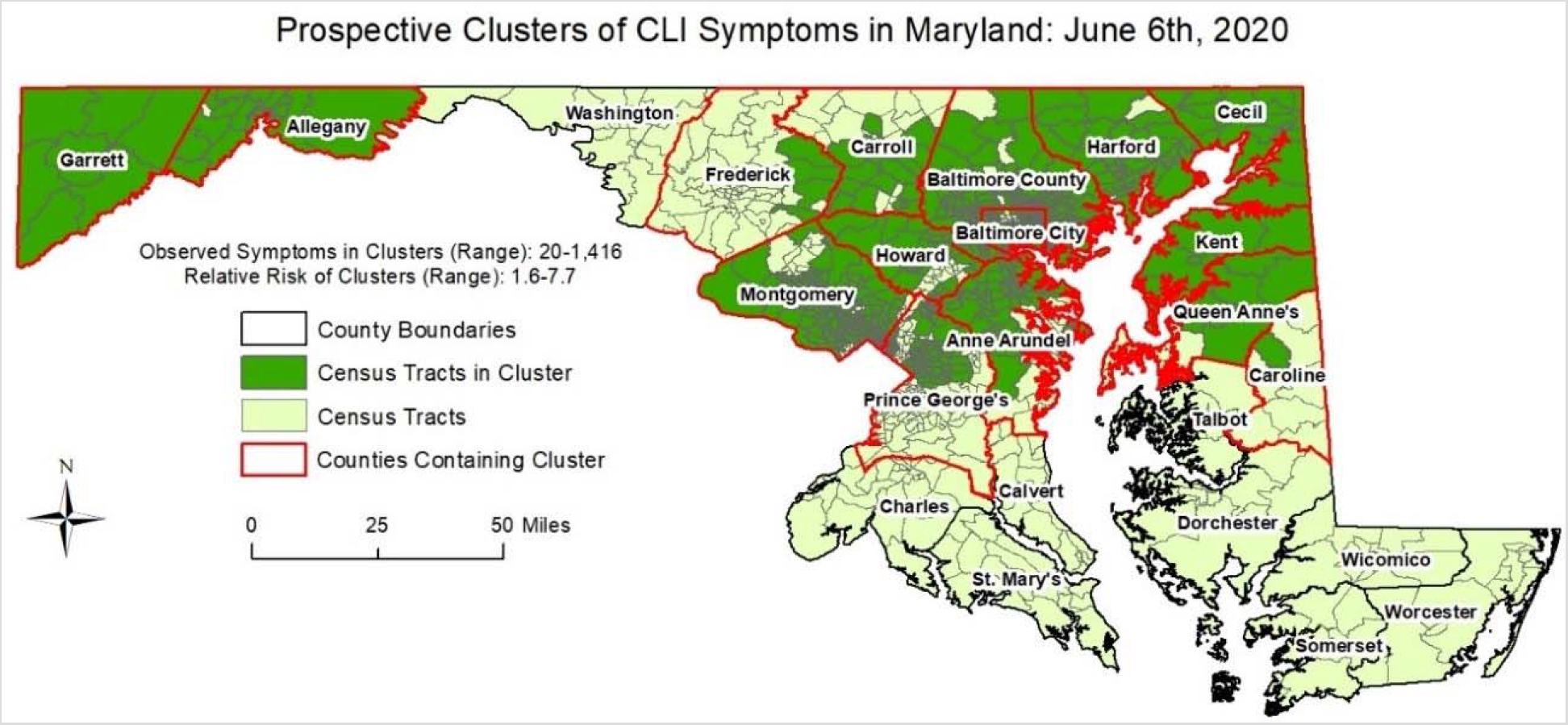
Space-time clusters of Simulated COVID Control Symptoms in Maryland on July 6^th^, 2020

**Figure 6.**
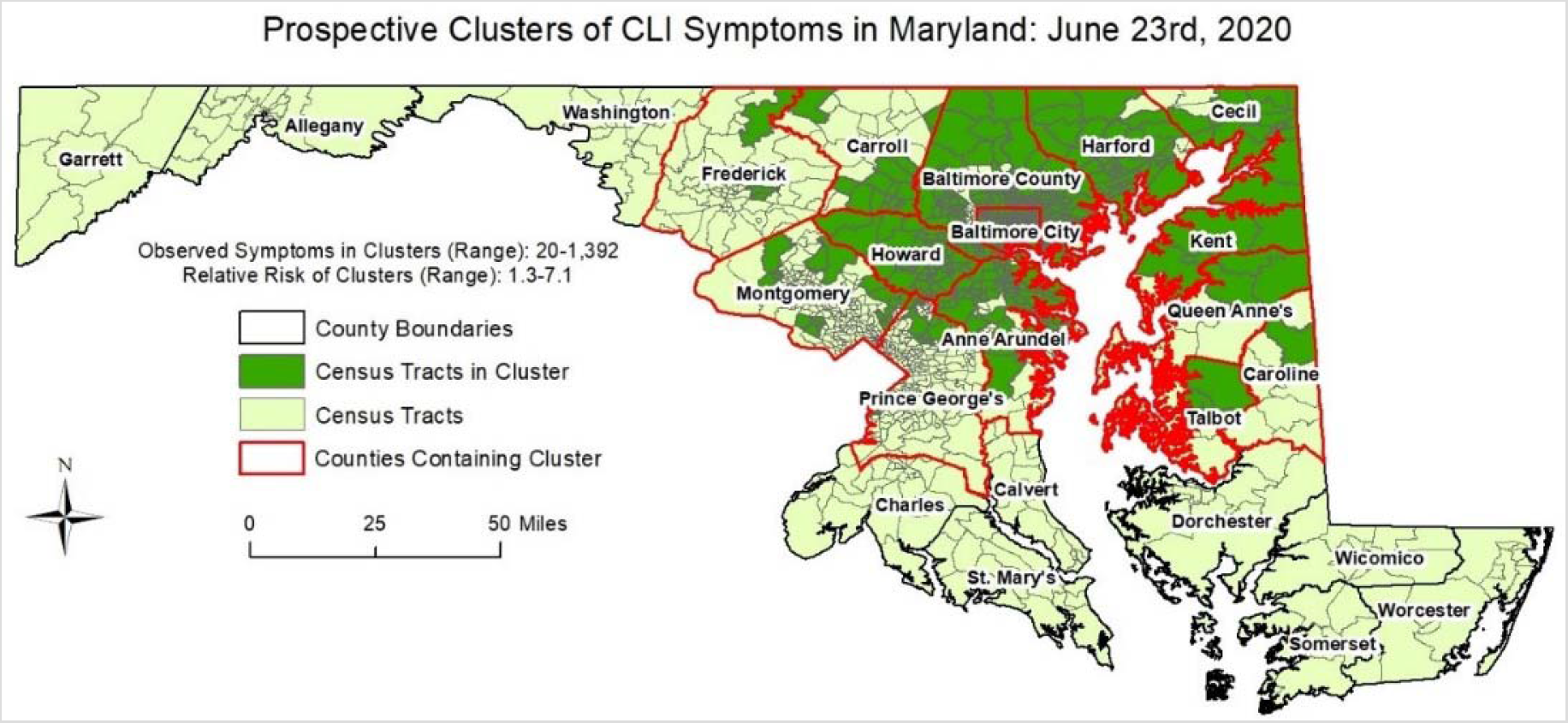
Space-time clusters of Simulated COVID Control Symptoms in Maryland on June 23^rd^, 2020

**Table 1.** Space-time cluster results using our simulated dataset in Maryland between May 26^th^ and June 23^rd^, 2020.

| County | Cluster duration (days) | Date cluster first identified | Last day cluster identified |
| --- | --- | --- | --- |
| Allegany | 9 | June 6<sup>th</sup> | June 14<sup>th</sup> |
| Anne Arundel | 22 | June 6<sup>th</sup> | June 23<sup>rd</sup> |
| Baltimore City | 22 | June 6<sup>th</sup> | June 23<sup>rd</sup> |
| Baltimore County | 22 | June 6<sup>th</sup> | June 23<sup>rd</sup> |
| Calvert | 0 | None | None |
| Caroline* | 12 | June 6<sup>th</sup>; June 23<sup>rd</sup> | June 17<sup>th</sup>; June 23<sup>rd</sup> |
| Carroll | 22 | June 6<sup>th</sup> | June 23<sup>rd</sup> |
| Cecil | 22 | June 6<sup>th</sup> | June 23<sup>rd</sup> |
| Charles | 0 | None | None |
| Dorchester | 0 | None | None |
| Frederick | 22 | June 6<sup>th</sup> | June 23<sup>rd</sup> |
| Garrett | 13 | June 6<sup>th</sup> | June 19<sup>th</sup> |
| Harford | 22 | June 6<sup>th</sup> | June 23<sup>rd</sup> |
| Howard | 22 | June 6<sup>th</sup> | June 23<sup>rd</sup> |
| Kent | 22 | June 6<sup>th</sup> | June 23<sup>rd</sup> |
| Montgomery | 22 | June 6<sup>th</sup> | June 23<sup>rd</sup> |
| Prince George's | 22 | June 6<sup>th</sup> | June 23<sup>rd</sup> |
| Queen Anne's | 22 | June 6<sup>th</sup> | June 23<sup>rd</sup> |
| Somerset | 0 | None | None |
| St. Mary's | 0 | None | None |
| Talbot | 1 | June 23<sup>rd</sup> | June 23<sup>rd</sup> |
| Washington | 0 | None | None |
| Wicomico | 0 | None | None |
| Worcester | 0 | None | None |

\*Caroline County exhibited no significant clusters between June 18<sup>th</sup> and June 22<sup>nd</sup>; then contained a space-time cluster of CLI-symptoms on June 23<sup>rd</sup>.

## Discussion

We have created a tool for large-scale syndromic surveillance that identifies geographical regions with abnormally high activity of fever and other COVID-19 like illness (CLI) symptoms, and preliminary results indicate that outbreaks of COVID-19 can be identified using this system. With validation, this system could be used to guide decision-making on health planning and resource allocation.

While several efforts have been implemented to monitor and predict COVID-19 cases or symptoms on a large scale (REFS), one of their biggest challenges is the approach for data collection. Some require a great effort on the part of users to complete long questionnaires. In others, questionnaires are nonspecific not allowing the user to feel engaged in the response to the pandemic. Both cases lead to the abandonment of citizen collaboration, which is essential in this situation. Moreover, these strategies provide results that are limited to county/state levels, but outbreaks typically do not stay contained within previously defined geographic areas. COVID Control app, unlike other apps with extensive surveys, presents a user-friendly and fast interface allowing users to submit relevant information in less than one minute. Moreover, users’ privacy is always guaranteed making it broadly accessible, as participants are not required to enter any personal information and automatic access to GPS location is optional. Finally, our analytics allow detection of clusters that are not restricted to pre-specified areas, making the results more comprehensive and realistic.

Acquiring data directly from individuals rather than hospitals/laboratories expands basic health monitoring of the general population and greatly reduces the delay in identifying new outbreaks of the disease. This, however, poses one of the major challenges because it relies on the engagement of the users to submit reliable self-reported information. For COVID Control to be effective, a large participation is required. Currently, spatial science and machine learning algorithms do a sufficient job in locating clusters but casting a wider net would certainly bolster the validity of the algorithms. Towards this objective, the strategies we are developing are two-fold. Firstly, efforts in advertising and creating awareness are needed to increase the number of new participants, especially in rural regions, and to get better resolution of data across the United states. Secondly, alongside more users, we need to increase sustained engagement of participants along time, including asymptomatic users’ CLI submissions. The latter is critical for a real-time and continuous monitoring of the health situation across the country, and to validate the models further. We found the majority of users (83.6%) self-report for a maximum of 14 days, the same time frame of common symptoms expression (see Figure 2).

With a larger userbase and further validation, this tool could be used to support a strategic response to prepare for an increase in hospitalizations and improved allocation of health care personnel and resources. Future work will implement a prospective cluster detection approach, which can detect “active” or emerging clusters of CLI symptoms, to give the most up-to-date public health overview.^18,19^ Since data in this report focused on Baltimore City and County in Maryland where we had the most users/entries, and COVID-19 data isn’t available at a resolution smaller than county, we have not validated the clusters’ location with confirmed case data. However, as the user base grows, validation work will be carried out. Cross-correlation analyses can also identify significant temporal lags between COVID-19 cases and symptoms collected from our app, which can improve upon our current analytical approach by informing the cluster analysis and validating our findings. In other words, we can potentially predict when and where COVID-19 outbreaks may arise in advance if we find a strong positive association between temporally lagged symptoms and confirmed COVID-19 cases. A strong positive association was already found in Baltimore City and County between new loss of taste/smell and positive cases of COVID-19. We expect that more data will increase the strength of associations between other symptoms, such as fever and other important COVID-19 related outcomes such as hospitalizations, helping to identify appropriate lags for each symptom-outcome pair which will subsequently improve the spatiotemporal cluster detection analysis.

Preemptive identification of potential clusters of COVID-19 can also be used by state and local authorities to develop a discriminative approach to reopening their economies guided by regionally specific syndromic information. The US government has included as a first criterion to be able to start the de-escalation plan to demonstrate a downward trajectory of flu and COVID-19 symptoms for at least 14 days, (taken from the guidelines that define the conditions that each state must meet before proceeding to the phased opening).^20^ Beyond the initial phase of reopening, this analysis would also potentially enable state and local governments to recalibrate their approach for specific counties / census tracts based on the observed trends. However, this type of approach for syndromic surveillance in isolation cannot address the issue of identifying asymptomatic carriers, but it can be used to optimize global testing strategies. COVID Control app and analytics are being continuously updated to consider the most recent discoveries and latest knowledge and integrate state-of-the-art approaches for identifying new cases as well as making it a useful tool to develop hypotheses of transmission. Finally, by immediately reporting the results of our analysis in a publicly available interactive dashboard, this tool allows citizens to actively participate in the surveillance of the pandemic, which increases their awareness of the disease and their response to it.^21^

## Methods

COVID Control: A Johns Hopkins University Study was approved by the Johns Hopkins School of Public Health Institutional Review Board (IRB number IRB00012283). The study has been performed in accordance with the institutional guidelines and regulations. All participants have voluntarily agreed to be involved in the study by accepting an IRB approved informed consent form when installing the app.

### COVID Control App Development

We developed an iOS and an Android app to gather symptom data from users. The app is available for free download from the Apple App Store and Google Play Store. Users are invited to voluntarily record their body temperature and, if applicable, the presence in the previous 24 hours of seven other symptoms that have been found to be good predictors of COVID-19: fever, cough, breathing difficulties, new loss of taste and/or smell, diarrhea, headache and fatigue.^11^ Among them, breathing difficulties, and new loss of taste and/or smell are more specific indicators of COVID-19.^11,22–24^ In addition, 88% of COVID-19 cases detected through surveillance and tests had a fever, making it a primary symptom for detecting the disease.^25^ A recent study also suggests body temperatures to be the most relevant determinant of contagions.^26^ The app users are instructed to use any available thermometer to measure and self-report their body temperature along with the observed symptoms.

The app does not record any individually identifiable health information and study participation is completely voluntary. Any individual above the age of 13 residing in the US is eligible for the study. To participate, users create an anonymous profile on the app by submitting their sex (‘Male’, ‘Female’, ‘Prefer not to say’) and age range (starting from 13 in increments of 5) (Figure 7). Information of the user location is collected at every submission. This can be via the phone’s GPS location (if access to location services is allowed) or by manual entry of the zip code by the user. All the information is assigned to a unique randomly generated ID and stored anonymously without any personal identification tags to a Microsoft Azure SQL database hosted on the cloud. When analyzed, the collected data is aggregated and combined with data from other users within the same county to contribute to the analytics and avoid any possible identification.

**Figure 7.**
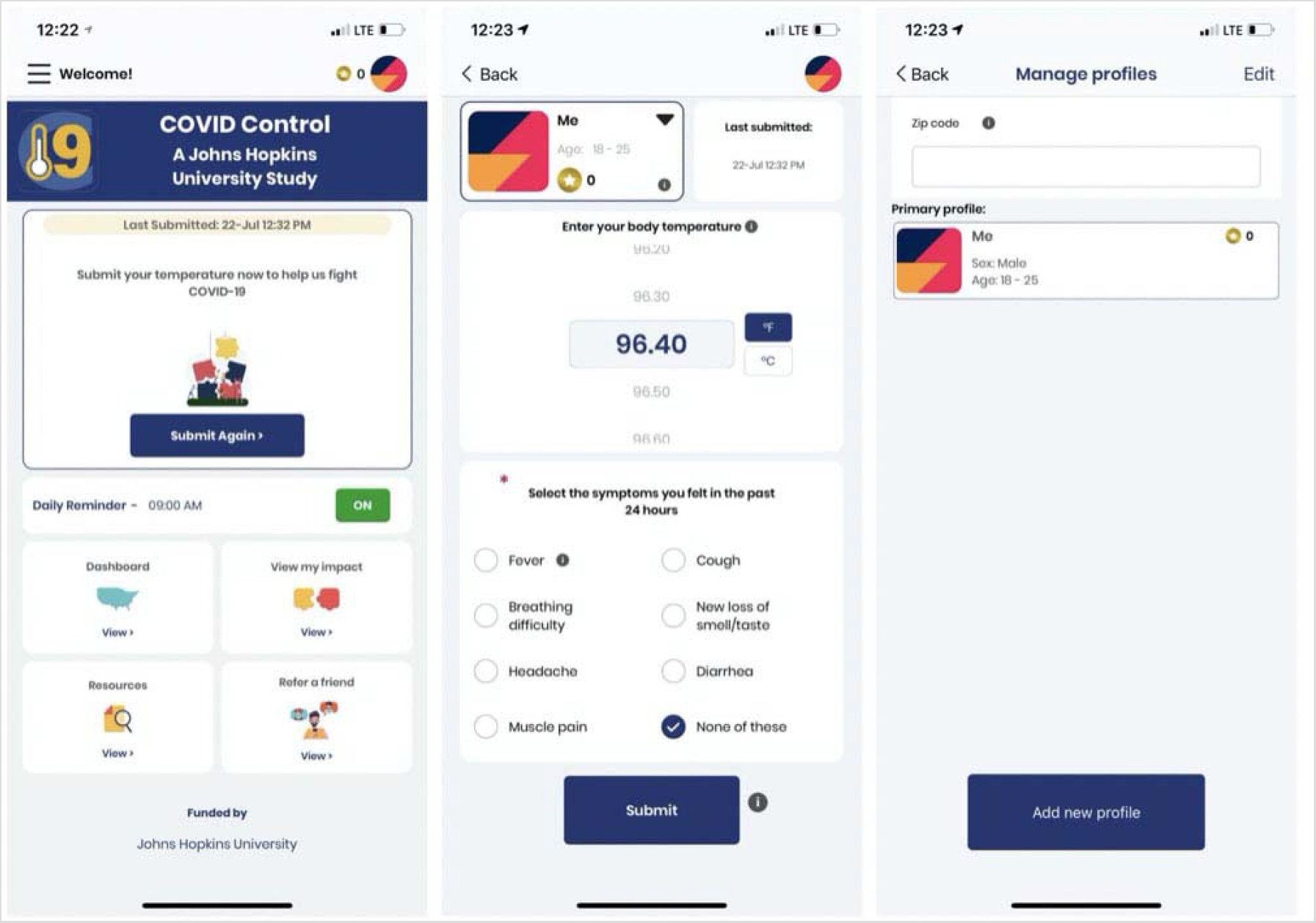
From to left to right: App main screen, app submission screen, and app profile screen.

The data collected through the app is aggregated by county and is presented back to users as an interactive dashboard, which provides a map of the symptom distribution, as well as time series plots of symptom rates. The dashboard allows users to focus on a specific geographic area, but also provides an overview for the entire United States. A link to the dashboard (Figure 8) embedded in the app allows the users to quickly visualize the dataset they are contributing to directly from their mobile device. A desktop version of the dashboard is also available on the App website both to app users as well as the public. The complete description of the data flow is depicted in Figure 9.

**Figure 8.**
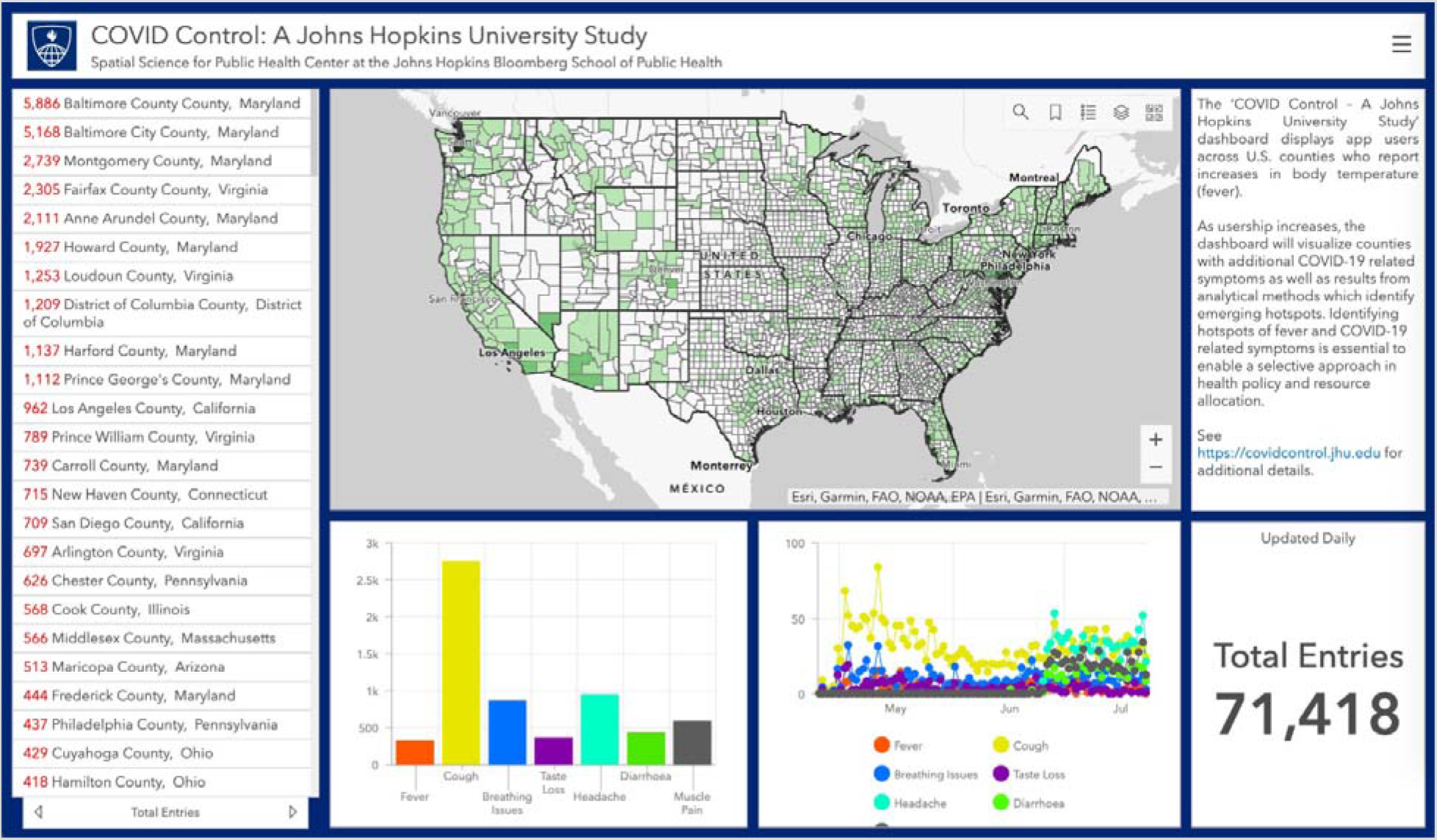
The dashboard provides an overview of submitted data across United States counties including entry and symptom numbers. Clicking on a county provides summary data for that specific area up to 23 June 2020.

**Figure 9.**
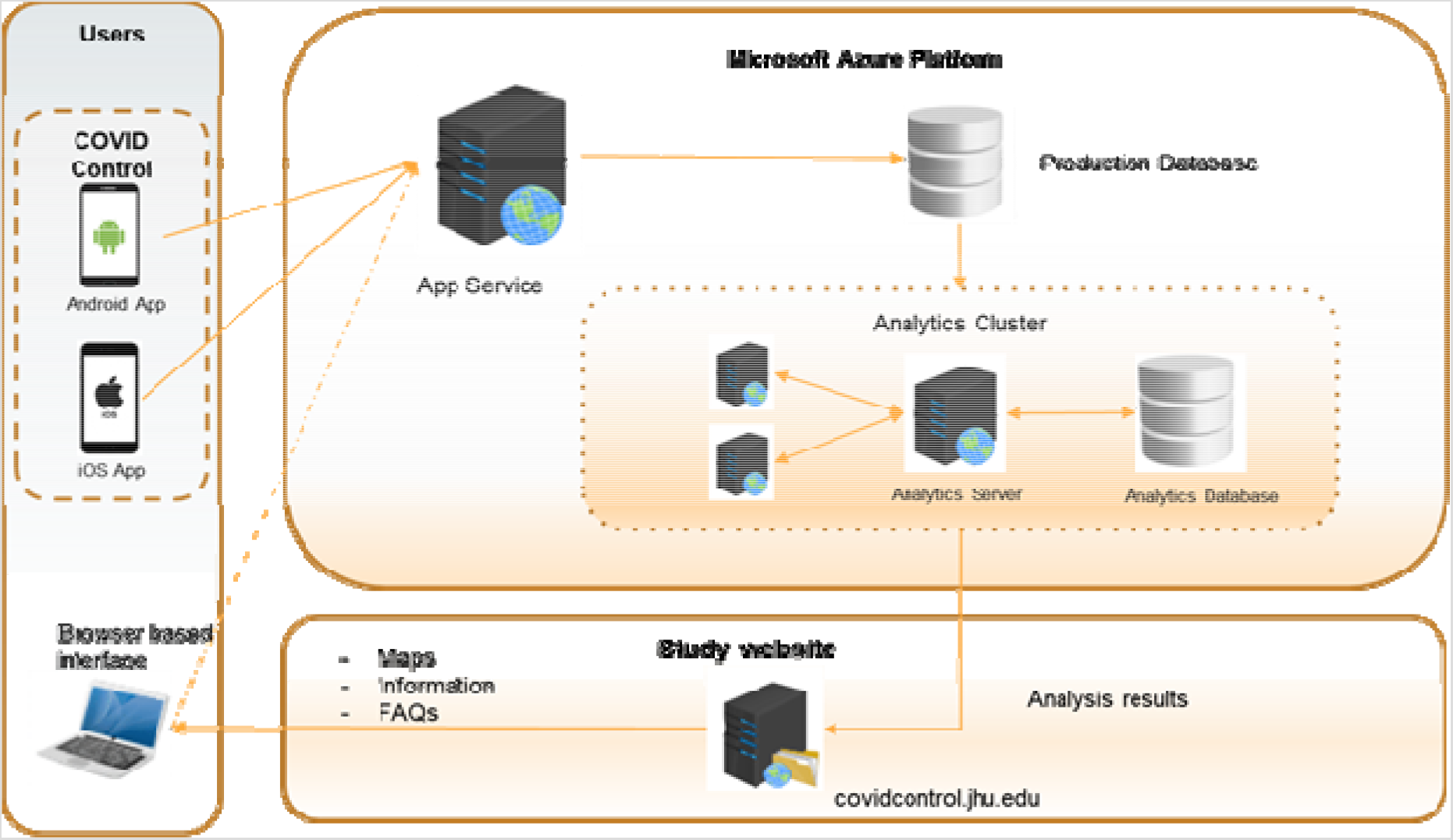
System design diagram representing the flow of data including the data collection from the apps, data storage and analysis in a Microsoft Azure server, and visualization of results in our Dashboard and website

### Cluster Detection Analysis

We utilize scan statistics, which are commonly used in epidemiology to detect and evaluate spatial, temporal, or spatiotemporal clustering of disease characteristics.^27,28^ Scan statistics are available using the free software SaTScan.^29^ SaTScan has been widely used in both disease and syndromic surveillance, including dengue fever and chikungunya,^17,30,31^ sexually transmitted disease;^32^ foodborne illness;^33^ respiratory infections and common illnesses;^34,35^ and COVID-19,^31,36,37^ among countless others. Essentially, scan statistics determine if the number of disease cases/symptoms in a defined area and proximal in time are greater than the expected number of cases/symptoms, such as the underlying population contained in the study area or distribution of point-level events. Our goal is to monitor the evolution of anomalous clusters of COVID-like illness (CLI) symptoms. We present an example at the daily and census-tract levels in Baltimore County and City and the U.S. state of Maryland. These locations were chosen since most users in the initial phase data collection phase are located in counties in close proximity to Johns Hopkins University.

We selected a retrospective approach and statistical model that detects significant historical clusters of CLI symptoms. The statistic utilizes circles (scanning window) that are centered on points (user locations with symptoms) and move (scan) systematically across a study area to identify clusters of symptoms (each window counts number of points within, while each scanning window is a potential cluster). Each scanning window is expanded in space to include neighboring points until a user-defined maximum radius is reached – here we selected 25 kilometers. Then, the number of observed symptoms within each window are compared to the expected number of symptoms. Before statistical inference is computed, a potential cluster is characterized when a scanning window contains more observed than expected symptoms. Space-time scan statistics^28^ incorporate a temporal dimension, where the scanning window is defined as a cylinder or three-dimensional ellipse, and the height represents the temporal dimension (e.g. time interval). The location, size, and duration of statistically significant clusters of disease cases are subsequently reported. Here, we use a retrospective, space-time permutation model. Monte Carlo simulation was used to compute statistical inference of the reported clusters of CLI symptoms; while all clusters are significant at the p < 0.05. To protect the privacy of the individuals who reported symptoms, we show the census tracts that belong to a significant cluster, rather than the spatial distribution of the points within each.

We also applied a cross-correlation function to identify the strongest association between each symptom available in the app and a variety of outcomes at the state-level in Maryland, including new positive tests, COVID-19 related deaths, and hospitalizations.^38^ The main purpose is to determine if clusters of symptoms may occur in advance of clusters of COVID-19 outcomes.

Finally, we utilized a prospective Bernoulli (i.e. cases/controls) version of the abovementioned space-time scan statistic on the simulated dataset to demonstrate how our analytics would work when our userbase increases. The model was run every day from June 6^th^ to June 23^rd^, 2020 to understand the space-time evolution of the detected clusters in Maryland. The maximum spatial extent for identified clusters was set to 2% of the population (i.e. simulated entries), while the maximum temporal extent for any identified cluster was set to 14 days. The mechanisms of the prospective model are essentially the same as the abovementioned retrospective model, except historical clusters are disregarded and only “active” and emerging clusters are reported on the most current day of analysis.

## Data Availability

The data may be available at the county level upon request.

## Acknowledgments

We thank John Rattray (Sparkwear) for his support developing the app, ITC Infotech’s Digital Experience (DX) team for further enhancing the app design, interface and UX, White and Case LLP for their guidance, Reina Murray and Mara Blake of the Department of Data Services of Johns Hopkins’ Sheridan Libraries and Museums for providing technical support for our dashboard, John Brown for his support with Microsoft Azure Server, Andrea Luxemberg (Johns Hopkins Medicine Technology Innovation Center) and Christian Tedjasukmana (University of Vermont Medical Center) for their contribution on user experience, and Jeff Trotz (Capitol Technology University). We would like to especially thank all the participants.

## Author Contributions Statement

The app development and data analytics and interpretation have been done by AG, SR, KA, MRD, AK, AC, BF, and TS. MS, AG, SR, KA have equally contributed to writing the manuscript. The study has been equally led and continuously supervised by FC, REC, and RS. All authors have contributed to the conception and design of the study and to the interpretation of results. All authors meet all four criteria for authorship defined in the ICMJE Recommendations.

## Ethics committee approval

COVID Control: A Johns Hopkins University Study was approved by the Johns Hopkins School of Public Health Institutional Review Board (IRB number IRB00012283). All participants have voluntarily agreed to be involved in the study by accepting an IRB approved consent form when installing the app.

